# An external, contemporary evaluation of the Epic End of Life Care Index among hospitalized patients across two large health systems: A retrospective cohort study

**DOI:** 10.64898/2026.01.02.26343350

**Authors:** Rachel Kohn, Katherine R. Courtright, Maria Grau-Sepulveda, Maren K. Olsen, Vanessa L. Madden, Bethany Sewell, Dorothy Sheu, Yazmeen S. Ahmad, Catherine L. Auriemma, Abigail Nimetz, Anne Dennos, Kimberly W. Hart, Janet Lee, Beth Creekmur, Claudia L. Nau, Huong Q. Nguyen, Susan Wang, Grant Boyer, Tammy Lundstrom, Lindsey Postema, Daniel J. Roth, James Vandewarker, Scott D. Halpern, Yuliya Lokhnygina

## Abstract

**Background:** The Epic End of Life Care Index (EOLCI) predicts one-year mortality and was developed to improve serious illness care. However, prior external EOLCI evaluations had limited sample sizes, populations, and equity evaluations. In preparation for a multi-system pragmatic clinical trial, we sought to evaluate the EOLCI performance and equity in the trial’s two participating health systems.

**Objective:** Evaluate EOLCI model performance overall and across key subgroups.

**Design/Setting/Patients:** Retrospective cohort study of patients hospitalized for ≥36 hours in 2022 to 39 hospitals in the Trinity Health and Kaiser Permanente Southern California (KPSC) health systems.

**Measurements:** We predicted one-year mortality risk stratified by health system using the EOLCI, a logistic regression model including age, sex, race/ethnicity, ethnicity, insurance, and diagnoses. We evaluated model performance using Scaled Brier Scores (SBS; range -1 to 1; composite measures of calibration and discrimination), calibration plots, and c-statistics.

**Results:** Among 116,749 Trinity patients with 154,063 encounters, 12,054 (10.3%) patients died within one year. Among 94,489 KPSC patients with 133,043 encounters, 16,872 (17.9%) died within one year. The SBS was -0.007 at Trinity and 0.178 at KPSC. Calibration was poor for both. Trinity’s discrimination was acceptable/good (c-statistic 0.76, 95% CI 0.76-0.77), and KPSC’s was good/very good (c-statistic 0.81, 95% CI 0.81-0.81). Model performance across subgroups was similar to the overall cohort.

**Limitations:** Death data were collected exclusively within Trinity and KPSC, risking outcome misclassification; several subgroup evaluations were limited by small sample sizes.

**Conclusions:** An external evaluation of the widely available Epic EOLCI demonstrated adequate to very good discrimination, poor calibration, and equitable performance across sociodemographic characteristics and diagnoses in two of the nation’s largest health systems.

**Primary funding source:** PCORI PLACER-2022C3-30553.

## INTRODUCTION

Palliative care (PC) is patient- and family-centered care that optimizes quality of life by anticipating, preventing, and treating suffering and burdens associated with a serious illness. Growing evidence consistently demonstrates that PC can improve the quality, experience, and outcomes of care,^1–3^ and it has been widely endorsed^4–9^ as a key strategy for health systems to improve the value of care.^10,11^ Thus, it is recommended that clinicians routinely address patients’ palliative needs themselves (generalist or primary PC) or consult medical experts to provide such care (specialist PC) at any time throughout the disease course.^4–9^ Despite this, many patients still never receive PC or only receive it in the final stages of life, limiting its potential benefits. To address this pervasive problem, health systems have increasingly deployed systematic approaches to identify patients most likely to benefit from PC.

‘Triggering PC,’ as it is often called, typically relies on a set of selected diagnostic, prognostic, or other clinical criteria available at the point of care. With the rapid growth of predictive analytics and health informatics, prognostic models based on electronic health record (EHR) data have become more commonplace for triggering PC.^12–14^ This approach is supported by evidence that PC improves the value of care for the sickest patients,^15–17^ and that prognostic uncertainty is a major barrier preventing clinicians from delivering timely PC.^18,19^ Some health systems have internally developed mortality risk models tailored to their unique patient populations, while many others rely on commercially available models.

Perhaps the most widely available commercial mortality risk model is the End of Life Care Index (EOLCI) developed by Epic. Epic is the most commonly used EHR system in United States hospitals, garnering nearly 40% of the EHR market share as of this writing. The EOLCI was specifically developed to identify adult patients who could benefit from advanced care planning, a fundamental component of generalist and specialist PC, by predicting one-year mortality risk. The EOLCI was trained and validated using EHR data from >550,000 patients across three health systems from 2014 through 2018. Per Epic’s EOLCI model brief, the final model demonstrated excellent discrimination on a low incidence outcome (i.e., overall one-year mortality), and poor calibration overall,^20^ and its performance was preserved across subgroups defined by sex, age, and race, and was superior among patients receiving Medicaid compared to commercial and Medicare recipients.

While recent studies have performed external evaluations of the EOLCI,^21–23^ generalizability of the findings are limited by inclusion of small sample sizes,^22,23^ few hospitals or clinics,^21–23^ and narrowly defined patient populations, such as older adults and patients with malignancy.^21–23^ In addition, these studies exclusively evaluated equity across sex, race, and ethnicity.^21–23^ Therefore, in preparation for a pragmatic comparative effectiveness trial that proposes to utilize the EOLCI to trigger clinician-directed nudges for PC in the EHR (NCT07224594),^24^ we sought to evaluate the EOLCI performance and equity across additional sociodemographic and diagnostic characteristics among a retrospective cohort of patients admitted to 39 hospitals across the two health systems participating in the trial.

## METHODS

### Study design and population

We performed a retrospective cohort study of all adult (≥18 years) inpatient encounters from patients admitted to 24 Trinity Health hospitals across eleven states and 15 Kaiser Permanente Southern California (KPSC) hospitals during 2022. The study cohort was restricted to encounters with a length of stay (LOS) ≥36 hours mimicking a key inclusion criterion for the parent trial. Vital status was obtained from the health systems with data collection ending December 31, 2023. We excluded patients admitted to psychiatry, acute rehabilitation, hospice, obstetrics, or neonatology inpatient services.

### Study variables

EOLCI predictive features included age, male sex, having Medicaid insurance, most recent albumin (g/dL) and red blood cell distribution width (RDW; %) within one year prior to admission through 36 hours after admission, the presence of comorbidities from encounters within one year prior to admission (International Classification of Diseases 10^th^ Revision diagnoses subsequently classified by the Healthcare Cost and Utilization Project Clinical Classifications Software [CCS])^25^ (eTable 1), and specific medication utilization (Table 1).^20^ Missing binary variables were presumed absent, and values were set at 0; missing lab values were given the median spline value; and age had no missing values.

**Table 1.** Encounter characteristics and model inputs – overall cohorts*.

| Characteristic | Trinity Health<br>n=154,063 | Kaiser Permanente Southern California<br>n=133,043 |
| --- | --- | --- |
| *Patient characteristic* |  |  |
| Age (years), median (IQR) | 69 (56-79) | 70 (57-80) |
| Age categories (years) |  |  |
| <50 | 25,682 (16.7%) | 21,769 (16.4%) |
| 50-64 | 36,558 (23.7%) | 29,042 (21.8%) |
| 65-74 | 37,218 (24.2%) | 31,587 (23.7%) |
| ≥75 | 54,605 (35.4%) | 50,645 (38.1%) |
| Female sex | 79,448 (51.6%) | 67,389 (50.7%) |
| Race |  |  |
| White | 115,941 (75.3%) | 79,888 (60.0%) |
| Black | 25,584 (16.6%) | 18,557 (13.9%) |
| Asian | 2,621 (1.7%) | 12,900 (9.7%) |
| Native American/Alaska Native | 505 (0.3%) | 668 (0.5%) |
| Native Hawaiian/Pacific Islander | 386 (0.3%) | 1,075 (0.8%) |
| Unknown | 3,658 (2.4%) | 15,700 (11.8%) |
| Other† | 3,293 (2.1%) | 4,255 (3.2%) |
| Declined | 2,037 (1.3%) | --- |
| Hispanic/Latinx ethnicity | 6,733 (4.4%) | 48,389 (36.4%) |
| Race/ethnicity categories |  |  |
| Non-Hispanic/Latinx white | 107,625 (69.9%) | 46,196 (34.7%) |
| Non-Hispanic/Latinx Black | 24,380 (15.8%) | 16,092 (12.1%) |
| Primary insurance |  |  |
| Private | --- | 45,381 (34.1%) |
| Medicare | 52,087 (33.8%) | 69,879 (52.5%) |
| Medicaid | 21,790 (14.1%) | 7,383 (5.5%) |
| Medicare and Medicaid | 20,615 (13.4%) | 9,988 (7.5%) |
| Self | 12,004 (7.8%) | --- |
| Other‡ | 47,567 (30.9%) | 412 (0.3%) |
| On Medicaid or both Medicare and Medicaid | 42,405 (27.5%) | 17,371 (13.1%) |
| EOLCI score, median (IQR) | 11% (4-27%) | 15% (4-42%) |
| Hospital length of stay (days), median (IQR) | 4.0 (2.6-6.7) | 3.4 (2.3-5.8) |
| One-year mortality (among 116,749 and 94,489 patients, respectively) | 12,054 (10.3%) | 16,872 (17.9%) |
| *Laboratory* |  |  |
| Albumin (g/dL), median (IQR) | 3.4 (3.0-3.9) | 3.1 (2.7-3.6) |
| Trinity: n=133,797 (86.8%)<br>KPSC: n=79,081 (59.4%) |  |  |
| RDW (%), median (IQR)<br>Trinity: n=152,646 (99.1%)<br>KPSC: n=132,829 (99.8%) | 14.4 (13.3-15.9) | 14.3 (13.3-15.9) |
| *Comorbidity* |  |  |
| Hepatitis | 2,655 (1.7%) | 3,378 (2.5%) |
| Cancer of head and neck | 771 (0.5%) | 1,248 (0.9%) |
| Cancer of bronchus; lung | 3,206 (2.1%) | 2,454 (1.8%) |
| Secondary malignancy | 4,197 (2.7%) | 9,054 (6.8%) |
| Maintenance chemotherapy; radiotherapy | 1,119 (0.7%) | 3,301 (2.5%) |
| Fluid and electrolyte disorder | 36,319 (23.6%) | 40,121 (30.2%) |
| Coagulation and hemorrhagic disorder | 9,224 (6.0%) | 15,254 (11.5%) |
| Parkinson's disease | 1,824 (1.2%) | 1,577 (1.2%) |
| Epilepsy; convulsions | 8,533 (5.5%) | 6,640 (5.0%) |
| Peri-; endo-; and myocarditis; cardiomyopathy | 10,724 (7.0%) | 9,453 (7.1%) |
| Coronary atherosclerosis and other heart diseases | 30,595 (19.9%) | 30,641 (23.0%) |
| Pulmonary heart disease | 12,214 (7.9%) | 7,003 (5.3%) |
| Conduction disorder | 14,676 (9.5%) | 12,169 (9.1%) |
| Heart failure; nonhypertensive | 31,070 (20.2%) | 32,554 (24.5%) |
| Acute cerebrovascular disease | 15,403 (10.0%) | 9,477 (7.1%) |
| Peripheral and visceral atherosclerosis | 14,344 (9.3%) | 64,384 (48.4%) |
| Chronic obstructive pulmonary disease and bronchiectasis | 25,117 (16.3%) | 18,792 (14.1%) |
| Respiratory failure; insufficiency; arrest (adult) | 24,188 (15.7%) | 22,879 (17.2%) |
| Other liver disease | 15,428 (10.0%) | 21,186 (15.9%) |
| Chronic ulcer of skin | 8,767 (5.7%) | 14,443 (10.9%) |
| Other injury or condition due to external causes | 18,194 (11.8%) | 17,269 (13.0%) |
| Nausea and vomiting | 11,529 (7.5%) | 16,105 (12.1%) |
| Delirium, dementia and amnestic and other cognitive disorders | 7,923 (5.1%) | 9,860 (7.4%) |
| Developmental disorders | 697 (0.5%) | 673 (0.5%) |
| Alcohol-related disorder | 10,078 (6.5%) | 8,848 (6.7%) |
| Screening and history of mental health and substance abuse | 6,716 (4.4%) | 8,358 (6.3%) |
| *Medication* |  |  |
| Albumin | 3,472 (2.3%) | 11,011 (8.3%) |
| Antiasthmatic | 60,344 (39.2%) | 46,877 (35.2%) |
| Antidiabetic | 52,249 (33.9%) | 57,133 (42.9%) |
| Antiemetic | 122,612 (79.6%) | 115,676 (86.9%) |
| Antineoplastic | 6,182 (4.0%) | 9,375 (7.0%) |
| Antipsychotic | 38,917 (25.3%) | 10,295 (7.7%) |
| Cardiotonic | 31,986 (20.8%) | 4,178 (3.1%) |
| Corticosteroid | 18,054 (11.7%) | 60,645 (45.6%) |
| Diuretic | 56,764 (36.8%) | 52,514 (39.5%) |
| Gastrointestinal prokinetic stimulant or agent | 8,035 (5.2%) | 53,079 (39.9%) |
| Laxative | 87,148 (56.6%) | 108,883 (81.8%) |
| Medical device | 16,681 (10.8%) | 48,548 (36.5%) |
| Opiate analgesic | 101,065 (65.6%) | 118,212 (88.9%) |
| Phosphate binder | 4,662 (3.0%) | 6,172 (4.6%) |
| Potassium removing resin | 1,348 (0.9%) | 1,250 (0.9%) |
| *Disease* |  |  |
| Advanced solid malignancy | 6,207 (4.0%) | 28,425 (21.4%) |
| Hematologic malignancy | 211 (0.1%) | 1,297 (1.0%) |
| Chronic obstructive pulmonary disease | 9,911 (6.4%) | 25,113 (18.9%) |
| Fibrotic (restrictive) lung disease | 318 (0.2%) | 5,738 (4.3%) |
| Liver disease | 1,872 (1.2%) | 11,792 (8.9%) |
| Neurodegenerative disease | 882 (0.6%) | 11,380 (8.6%) |
| Heart failure | 19,526 (12.7%) | 49,364 (37.1%) |
| Chronic kidney disease | 5,452 (3.5%) | 21,823 (16.4%) |
| **Abbreviations:** RDW, red blood cell distribution width; EOLCI, Epic End of Life Care Index; IQR, interquartile range; KPSC, Kaiser Permanente Southern California<br>*Variables reported as n (%) unless otherwise indicated<br>†Other includes: Middle Eastern/North African; multiracial; other<br>‡Other includes: private (Trinity Health); other (both health systems) |  |  |

The outcome was one-year mortality, measured using all data available within each health system’s EHR, commencing at hour 36 of included hospital admissions.

### Analyses

For the EOLCI, age, albumin, and RDW were included in the model as continuous variables. The remainder were binary. EOLCI scores were generated within each health system, ranging from 0-100%.

We evaluated model performance overall and by subgroup within each health system using Scaled Brier Scores (SBS; range -1 to 1, higher scores are better; composite measures of calibration and discrimination that incorporate the difference between observed and expected outcomes, number of observations, and event rates);^26^ calibration plots (agreement between predicted probabilities [i.e., EOLCI scores] of predicted vs observed one-year mortality from 36 hours after admission); and discrimination using receiver operating characteristic (ROC) curves and area under the ROC curve (AUC), also known as c-statistics (>0.90 was considered excellent; 0.80-0.89 good/very good; 0.70-0.79 acceptable/good; 0.65-0.69 weak/acceptable; 0.60-0.64 poor; 0.50-0.59 unacceptable). We additionally evaluated true positives, false positives, true negatives, false negatives, positive predictive values (PPV), negative predictive values (NPV), sensitivity, specificity, false positive rates (FPR), and false negative rates (FNR) at specific thresholds of the EOLCI.

To generate our calibration curves, we divided our populations into deciles of predicted probabilities and plotted the average predicted probability against the observed mortality risk in each decile. We computed confidence intervals (CI) using the exact binomial formula. To calculate c-statistics, we used logistic regression, treating the observed vital status (0 for alive, 1 for dead) as the dependent variable and the EOLCI one-year predicted mortality risk as the independent variable. We forced the predicted mortality risk coefficient to be 1 to prevent potential bias introduced by the scaling of predicted probabilities.^27^ This approach allowed us to directly assess the discriminatory power of the predicted probabilities in ranking individuals according to their mortality risk in the evaluation dataset.

### Subgroups

We specified subgroup populations a priori within which to evaluate equity: age (<50, 50-65, >65-75, >75 years), sex (female, male), race/ethnicity (non-Hispanic Black, non-Hispanic white), ethnicity (Hispanic/Latinx, not Hispanic/Latinx), insurance (Medicaid or dual-eligible, non-Medicaid or non-dual-eligible), and diagnoses (eTable 2). We evaluated performance metrics of the EOLCI at risk thresholds ranging from 50% to 80% in 10% increments and from 85% to 95% in 5% increments.

All analyses were performed in SAS, version 9.4 (Cary, NC). The study was approved by the University of Pennsylvania Institutional Review Board.

## RESULTS

Results are reported following STROBE guidelines.

### General characteristics

Within Trinity hospitals, 116,749 patients had 154,063 encounters. Across all encounters, median age was 69 years (interquartile range [IQR] 56-79 years), 79,448 (51.6%) were female, 115,941 (75.3%) were white, 25,584 (16.6%) were Black, 6,733 (4.4%) were Hispanic/Latinx, and 42,405 (27.5%) were primarily insured by Medicaid or both Medicare and Medicaid. 6,207 (4.0%) patients had advanced solid malignancies, 211 (0.1%) had hematologic malignancies, 9,911 (6.4%) had chronic obstructive pulmonary disease (COPD), 19,526 (12.7%) had heart failure, and 5,452 (3.5%) had chronic kidney disease (CKD). The median EOLCI score was 11% (IQR 4-27%). Median hospital LOS was 4.0 days (IQR 2.6-6.7), and 12,054 (10.3%) patients died within one year of the 36^th^ hour of their hospitalization.

Within KPSC hospitals, 94,489 patients had 133,043 encounters. Across all encounters, median age was 70 years (IQR 57-80 years), 67,389 (50.7%) were female, 79,888 (60.0%) were white, 18,557 (13.9%) were Black, 48,389 (36.4%) were Hispanic/Latinx, and 17,371 (13.1%) were primarily insured by Medicaid or both Medicare and Medicaid. 28,425 (21.4%) patients had advanced solid malignancies, 1,297 (1.0%) had hematologic malignancies, 25,113 (18.9%) had COPD, 49,364 (37.1%) had heart failure, and 21,823 (16.4%) had CKD. The median EOLCI score was 15% (IQR 4-42%). Median hospital LOS was 3.4 days (IQR 2.3-5.8), and 16,872 (17.9%) patients died within one year of the 36^th^ hour of their hospitalization (Table 1).

### Overall model performance of the Epic End of Life Care Index (EOLCI)

The overall SBS was -0.007 across Trinity encounters and 0.178 across KPSC encounters, indicating poor and moderate composite measures of calibration and discrimination across Trinity and KPSC encounters, respectively. Calibration was overall poor in both health systems. In Trinity, for observed one-year risk of mortality from the 36^th^ hour of hospitalization >8%, the EOLCI model overpredicted mortality. In KPSC, calibration was good for observed one-year risk of mortality from the 36^th^ hour of hospitalization <30%, but for observed mortality risk >30%, the EOLCI model overpredicted mortality (Figure 1). Discrimination was acceptable/good in Trinity (c-statistic 0.76, 95% CI 0.76-0.77), and good/very good in KPSC (c-statistic 0.81, 95% CI 0.81-0.81) (Figure 2).

**Figure 1.**
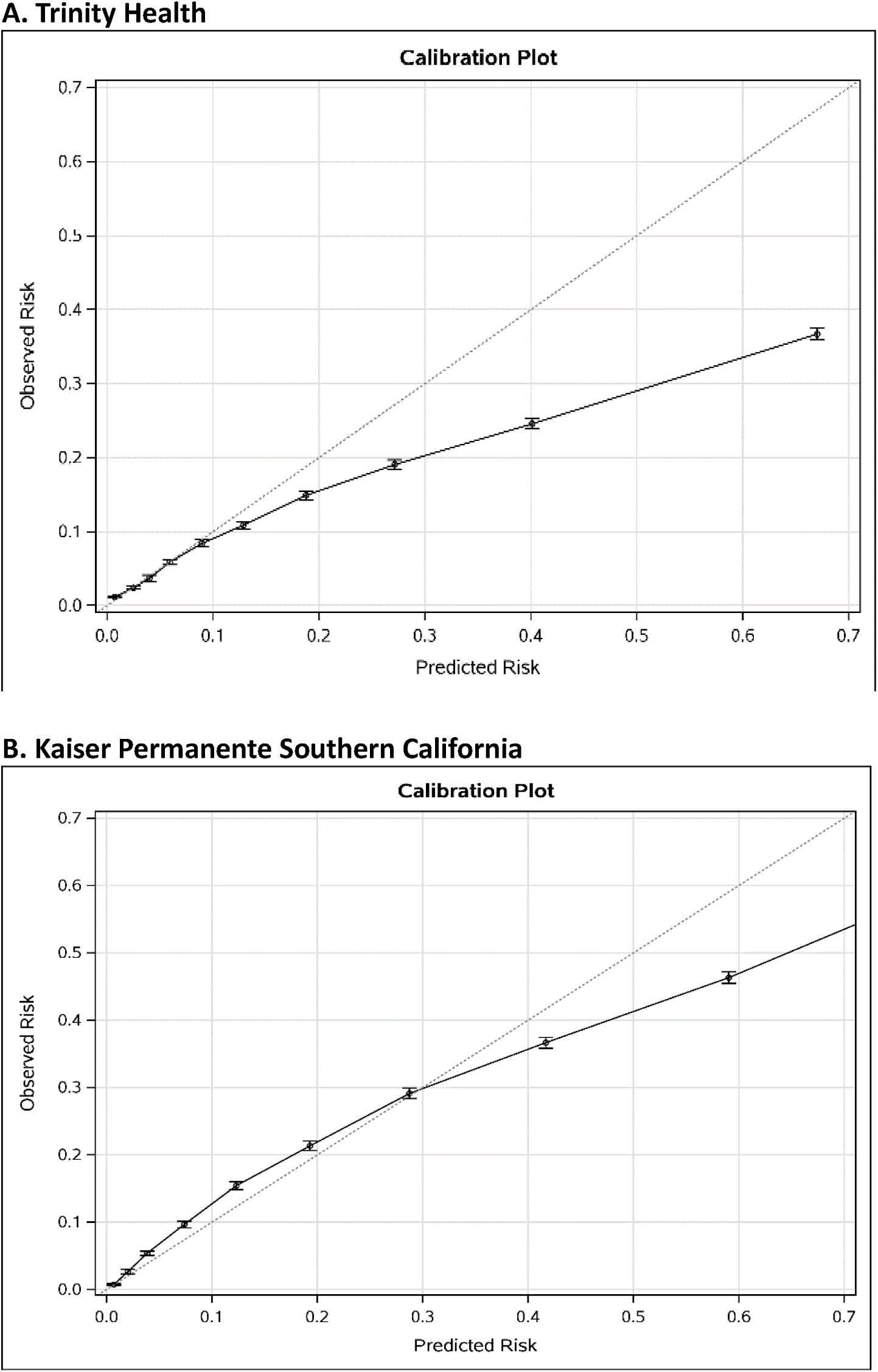
Calibration plots of Epic End of Life Care Index predicted probabilities of 1-year mortality from the 36^th^ hour of hospitalization vs observed mortality for A. Trinity Health and B. Kaiser Permanente Southern California (KPSC)

**Figure 2.**
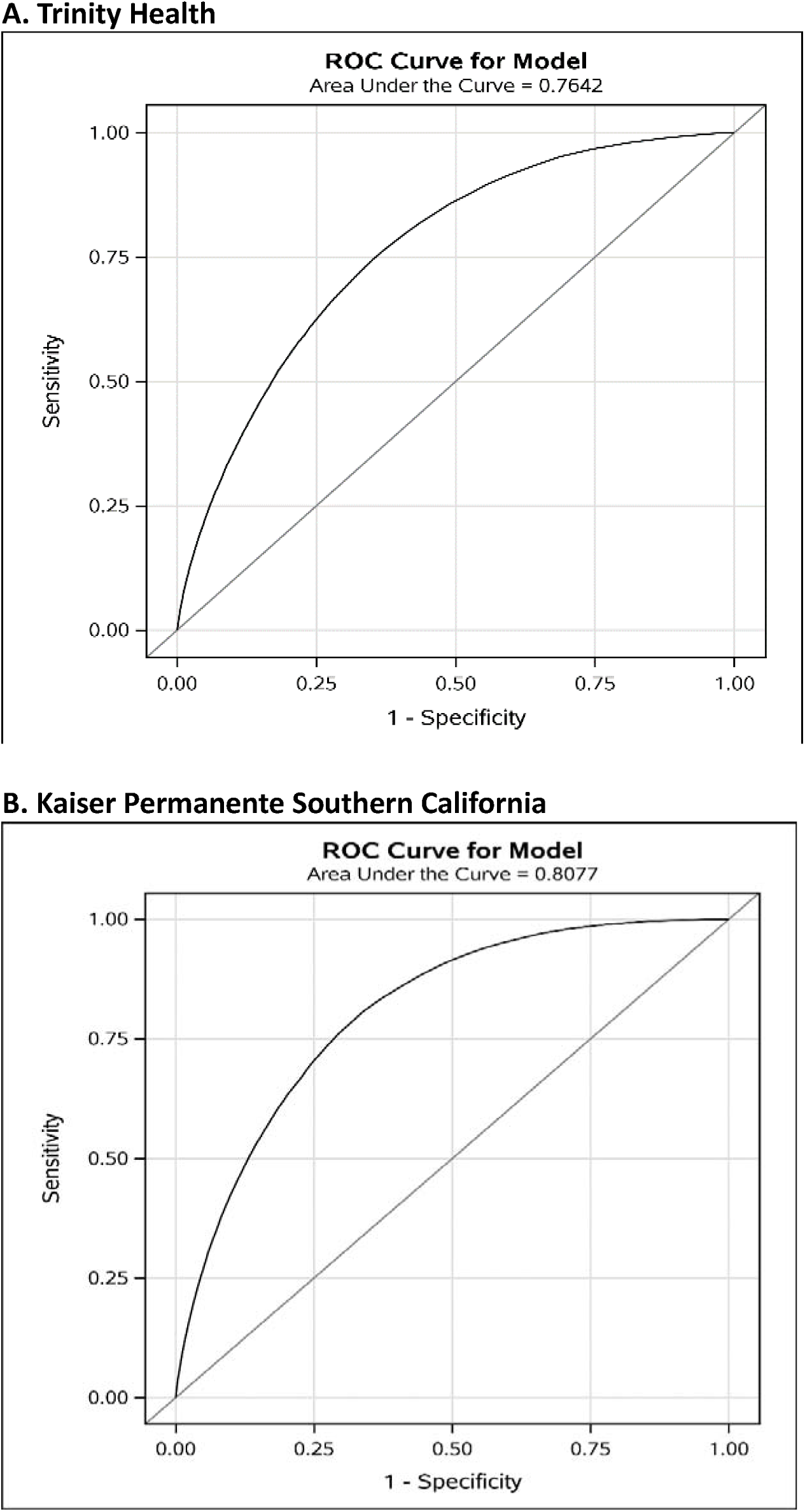
Receiver operating characteristic (ROC) curves of Epic End of Life Care Index predicted probabilities of 1-year mortality from the 36^th^ hour of hospitalization for A. Trinity Health and B. Kaiser Permanente Southern California

In Trinity, risk-group-specific true positives ranged from 198-5,681 for EOLCI risk score thresholds of 95% to 50%, respectively. Risk-group-specific PPVs ranged from 0.367-0.529, specificity from 0.927-0.999, and FNRs from 0.712-0.990 for EOLCI risk score thresholds of 50% to 95%, respectively. Risk-group-specific NPVs ranged from 0.873-0.899, sensitivity from 0.010-0.288, and FPRs from 0.001-0.073 for EOLCI risk score thresholds of 95% to 50%, respectively. At the EOLCI threshold of 70% one-year mortality risk, our proposed threshold for intervention within our pragmatic comparative effectiveness trial, true positives were 2,640, PPV was 0.445, NPV 0.885, sensitivity 0.134, specificity 0.976, FPR 0.024, and FNR 0.866 (Table 2A, Figure 3A).

**Figure 3.**
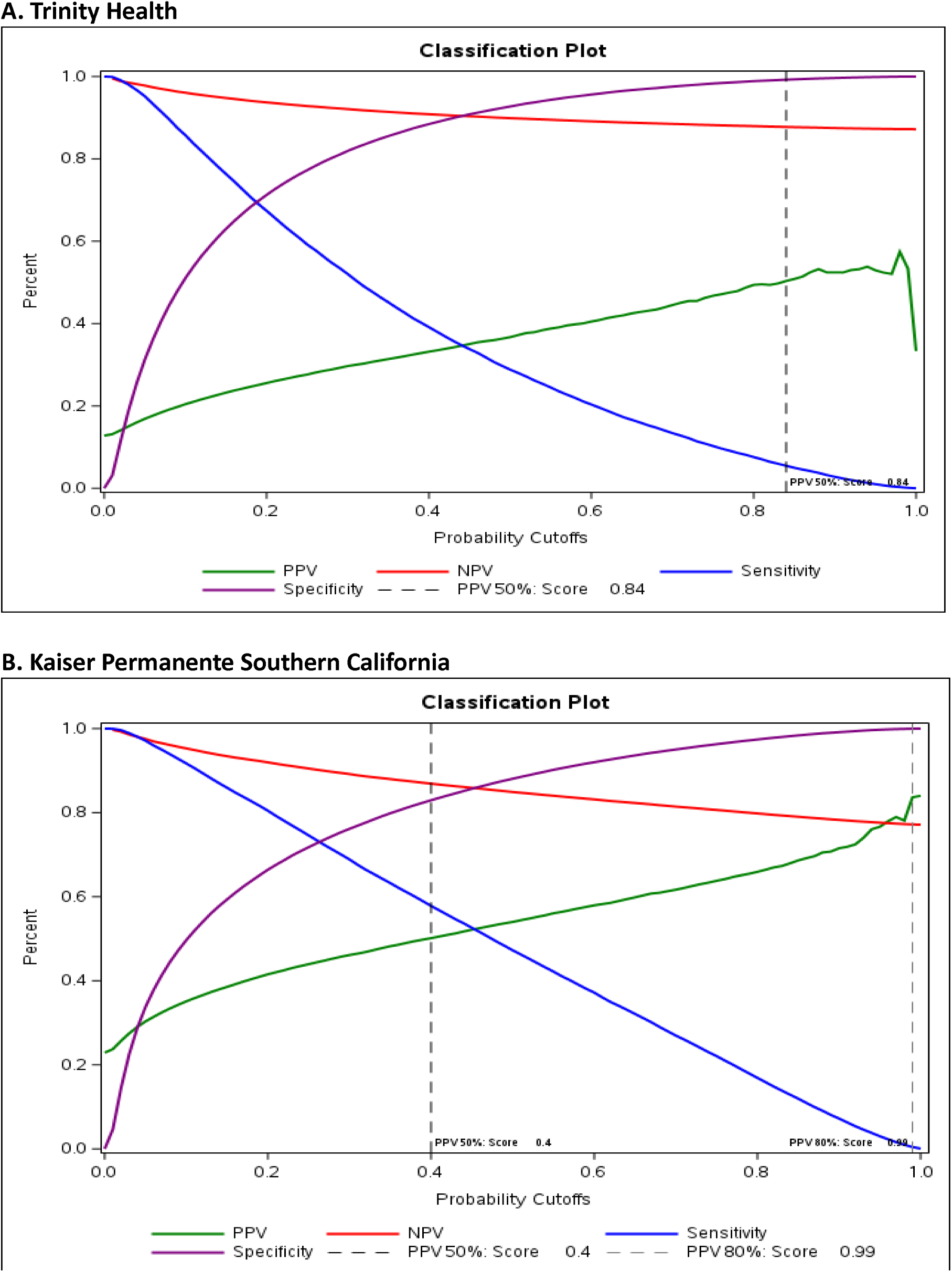
Relationship of Epic End of Life Care Index performance metrics for A. Trinity Health and B. Kaiser Permanente Southern California

**Table 2.** Classification tables of Epic End of Life Care Index (EOLCI) performance metrics for A. Trinity Health and B. Kaiser Permanente Southern California.

**A. Trinity Health**
| EOLCI score thresholds | True positives | False positives | True negatives | False negatives | Positive predictive value | Negative predictive value | Sensitivity | Specificity | False positive rate | False negative rate |
| --- | --- | --- | --- | --- | --- | --- | --- | --- | --- | --- |
| 50% | 5,681 | 9,797 | 124,558 | 14,027 | 0.367 | 0.899 | 0.288 | 0.927 | 0.073 | 0.712 |
| 60% | 4,012 | 5,895 | 128,460 | 15,696 | 0.405 | 0.891 | 0.204 | 0.956 | 0.044 | 0.796 |
| 70% | 2,640 | 3,291 | 131,064 | 17,068 | 0.445 | 0.885 | 0.134 | 0.976 | 0.024 | 0.866 |
| 80% | 1,495 | 1,531 | 132,824 | 18,213 | 0.494 | 0.879 | 0.076 | 0.989 | 0.011 | 0.924 |
| 85% | 981 | 949 | 133,406 | 18,727 | 0.508 | 0.877 | 0.050 | 0.993 | 0.007 | 0.950 |
| 90% | 551 | 500 | 133,855 | 19,157 | 0.524 | 0.874 | 0.028 | 0.996 | 0.004 | 0.972 |
| 95% | 198 | 176 | 134,179 | 19,510 | 0.529 | 0.873 | 0.010 | 0.999 | 0.001 | 0.990 |

| EOLCI score thresholds | True positives | False positives | True negatives | False negatives | Positive predictive value | Negative predictive value | Sensitivity | Specificity | False positive rate | False negative rate |
| --- | --- | --- | --- | --- | --- | --- | --- | --- | --- | --- |
| 50% | 14,381 | 12,254 | 90,348 | 16,060 | 0.540 | 0.849 | 0.472 | 0.881 | 0.119 | 0.528 |
| 60% | 11,317 | 8,212 | 94,390 | 19,124 | 0.580 | 0.832 | 0.372 | 0.920 | 0.080 | 0.628 |
| 70% | 8,202 | 5,098 | 97,504 | 22,239 | 0.617 | 0.814 | 0.269 | 0.950 | 0.050 | 0.731 |
| 80% | 5,140 | 2,658 | 99,944 | 25,301 | 0.659 | 0.798 | 0.169 | 0.974 | 0.026 | 0.831 |
| 85% | 3,636 | 1,662 | 100,940 | 26,805 | 0.686 | 0.790 | 0.119 | 0.984 | 0.016 | 0.881 |
| 90% | 2,197 | 875 | 101,727 | 28,244 | 0.715 | 0.783 | 0.072 | 0.991 | 0.009 | 0.928 |
| 95% | 915 | 279 | 102,323 | 29,526 | 0.766 | 0.776 | 0.030 | 0.997 | 0.003 | 0.970 |

In KPSC, risk-group-specific true positives ranged from 915-14,381 for EOLCI risk score thresholds of 95% to 50%, respectively. Risk-group-specific PPVs ranged from 0.540-0.766, specificity from 0.881-0.997, and FNRs from 0.528-0.970 for EOLCI risk score thresholds of 50% to 95%, respectively. Risk-group-specific NPVs ranged from 0.776-0.849, sensitivity from 0.030-0.472, and FPRs from 0.003-0.119 for EOLCI risk score thresholds of 95% to 50%, respectively. At the EOLCI threshold of 70%, true positives were 8,202, PPV was 0.617, NPV 0.814, sensitivity 0.269, specificity 0.950, FPR 0.050, and FNR 0.731 (Table 2B, Figure 3B).

### Subgroup model performance of the Epic End of Life Care Index (EOLCI)

Across both Trinity and KPSC, the model performed similarly in pre-specified subgroups as in the overall model for SBS and c-statistics, with the exception that in both health systems, increasing age was associated with worse SBS, c-statistics, NPVs, FPRs, and specificity, and with increased sensitivity and FNRs. Performance characteristics were similar across all diagnostic categories at the chosen EOLCI threshold of 70% mortality risk (Table 3).

**Table 3.**
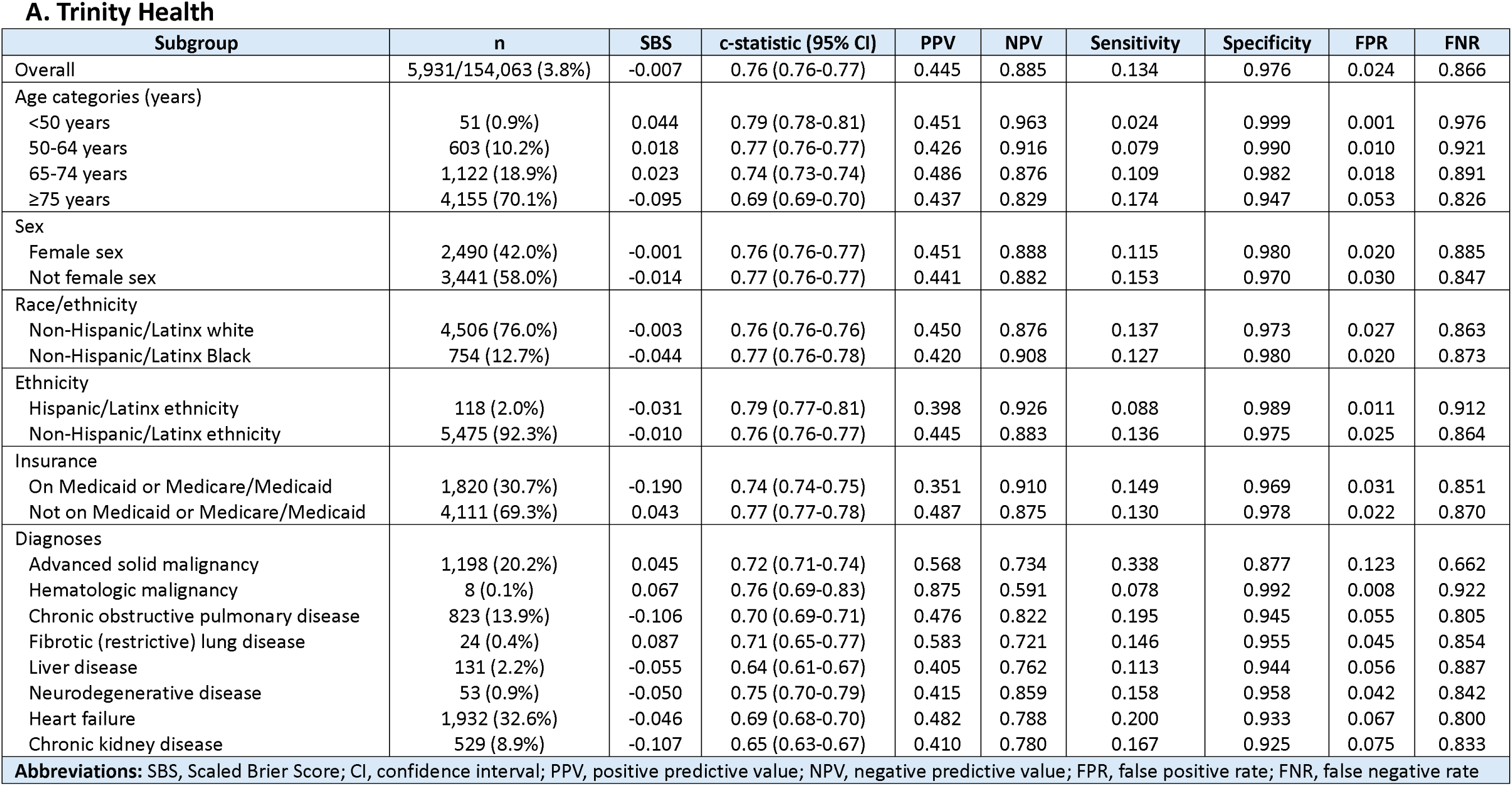
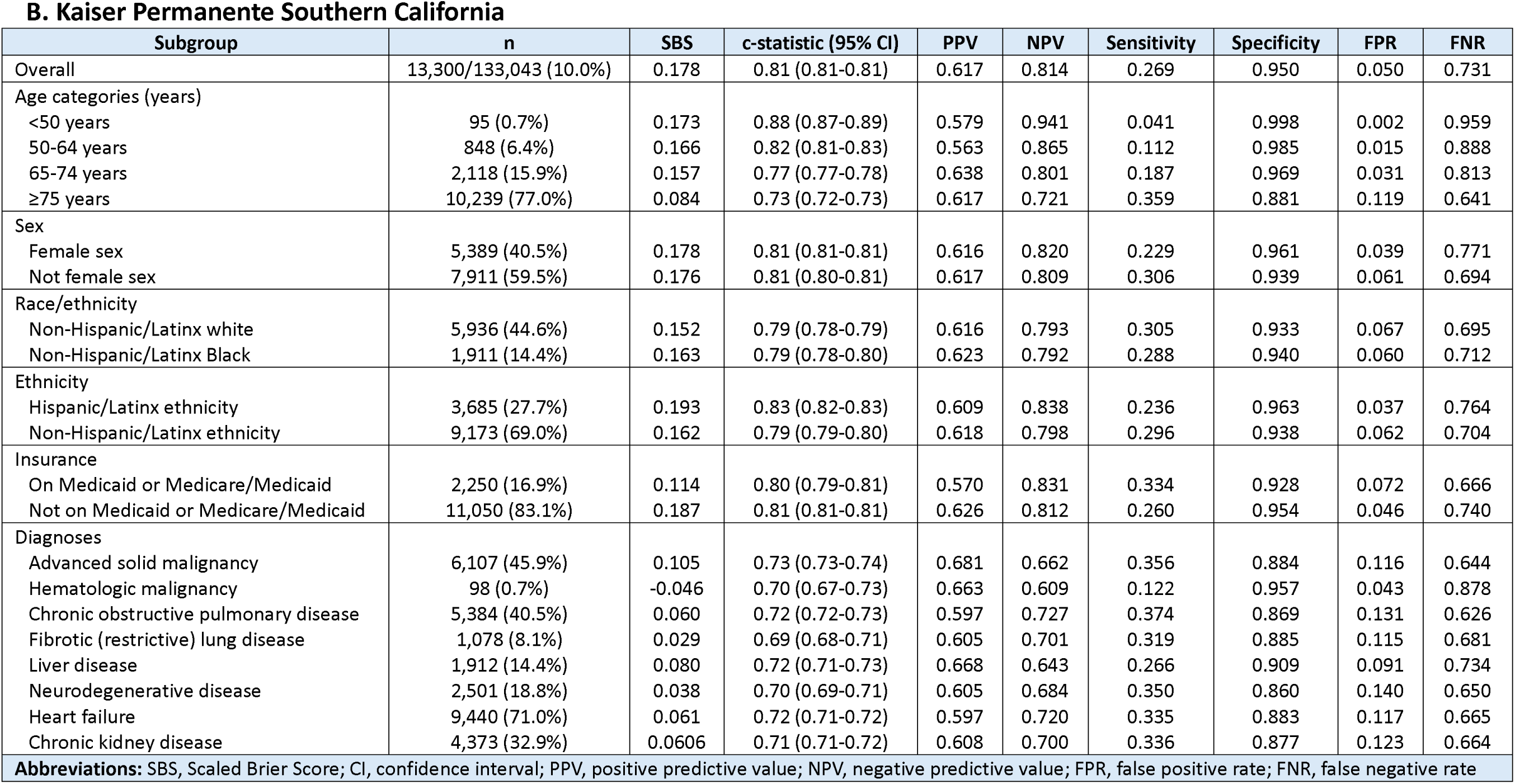
Epic End of Life Care Index performance metrics by subgroup at a predicted mortality risk of 70% for A. Trinity Health and B. Kaiser Permanente Southern California.

## DISCUSSION

This study represents the largest, publicly available external evaluation of the Epic EOLCI, including 13 times more hospitals than the model development cohort and >6 times more hospitals than the next largest external evaluation.^21^ The original EOLCI found c-statistics of 0.89-0.90, and subsequent evaluations of the EOLCI have found c-statistics ranging from 0.71-0.82, and that performance was preserved across subgroups defined by age, sex, and race.^20–23^ We have both replicated these findings and expanded the equity evaluation to include Hispanic/Latinx ethnicity, insurance, and diagnoses common among patients with serious illnesses. In doing so, we demonstrated that the EOLCI has adequate to very good discrimination, poor calibration, and performs equitably across sociodemographic and diagnostic characteristics in 39 hospitals across two of the nation’s largest health systems.

These findings are important given the vast number of hospitals already using the EOLCI, and the hundreds of other U.S. hospitals with the ability to use it, particularly those without the resources to develop de novo local risk models. Our findings suggest the model is suitable for prospectively differentiating among hospitalized patients who will and will not die in the following year, which could inform implementation and utilization for myriad clinical use cases. Beyond screening patients to identify those who may benefit from PC and advance care planning or goals of care discussions, the EOLCI may be useful for directing resource allocation, such as case management or social services, or screening patients for participation in research studies. However, calibration was overall poor in both health systems, and particularly poor among older age groups. The EOLCI may therefore not be suitable for use cases such as prognostic communication, determining hospice eligibility, or risk adjustment in retrospective research. As such, health systems may consider recalibrating the model to their specific patient populations prior to implementation and use for these and other similar use cases. In addition, true positives varied by magnitudes depending on the EOLCI score thresholds utilized, to as low as 198 and 915 patients in Trinity and KPSC, respectively, using a 95% threshold. Thus, risk threshold selection must be context-specific. While a 95% threshold may be appropriate in a clinical trial setting for predictive or prognostic enrichment, for example, it may not be an appropriate threshold for a hospital-wide quality improvement initiative. For these reasons, all EOLCI implementation decisions should be made at the individual health system level based on local context.

Additionally, in evaluating whether to implement a model, it is crucial to consider external model performance metrics. The EOLCI was developed and validated in 2014-2018 data among >550,000 patients across three hospitals, and demonstrated c-statistics of 0.89-0.90, PPVs of 0.30-0.55, and sensitivities of 0.05-0.10 at score thresholds of 70% mortality risk.^20^ In our study, in the overall population, the c-statistic was 0.76 for Trinity and 0.81 for KPSC. At EOLCI risk score thresholds of 70% mortality risk, PPVs were 0.45 and 0.62, and sensitivities were 0.13 and 0.27 for Trinity and KPSC, respectively. This demonstrates that the EOLCI discrimination performs worse in our external evaluation, while the PPV and sensitivity were stable to improved.

Our study has several limitations. First, we could only identify deaths if they were recorded in the participating health systems’ EHRs. With KPSC being an integrated health system, this concern is largely mitigated, but we may nonetheless have misclassified patients as alive if deaths occurred outside the health system. Second, we were unable to evaluate performance equity by gender identity or races other than white or Black, given the small sample sizes in those subgroups. Third, race and ethnicity were derived from the ‘race/ethnicity’ field in the EHR, which may be inaccurate due to a lack of fidelity in the data gathering process.^28,29^ Fourth, as with all studies using EHR data, patient characteristics and comorbidities may be underdiagnosed, misclassified, or underreported.

In conclusion, this contemporary external evaluation of the Epic EOLCI in 39 diverse U.S. hospitals within two large health systems demonstrated adequate to very good discrimination, poor calibration, and equitable performance across sociodemographic and diagnostic characteristics. These findings may inform new opportunities for the appropriate use of the EOLCI in clinical settings and serve as a benchmark for the development of local models predicting one-year mortality.

## Supporting information

eTable 1, eTable 2

## Data Availability

All data produced in the present study are available upon reasonable request to the authors

## Funding acknowledgement

This work was supported through a Patient-Centered Outcomes Research Institute (PCORI) Award (PLACER-2022C3-30553)

