## Supplementary material for "An external, contemporary evaluation of the Epic End of Life Care Index among hospitalized patients across two large health systems: A retrospective cohort study": eTable 1, eTable 2

**SUPPLEMENT**

**SUPPLEMENTAL TABLES**

**eTable 1. Epic End of Life Care Index comorbidities**

| **Diagnosis** | **Clinical Classifications Software group** |
| --- | --- |
| Hepatitis | CCS 6 |
| Cancer of head and neck | CCS 11 |
| Cancer of bronchus; lung | CCS 19 |
| Secondary malignancy | CCS 42 |
| Maintenance chemotherapy; radiotherapy | CCS 45 |
| Fluid and electrolyte disorder | CCS 55 |
| Coagulation and hemorrhagic disorder | CCS 62 |
| Parkinson’s disease | CCS 79 |
| Epilepsy; convulsions | CCS 83 |
| Peri-; endo-; and myocarditis; cardiomyopathy | CCS 97 |
| Coronary atherosclerosis and other heart diseases | CCS 101 |
| Pulmonary heart disease | CCS 103 |
| Conduction disorder | CCS 105 |
| Heart failure; nonhypertensive | CCS 108 |
| Acute cerebrovascular disease | CCS 109 |
| Peripheral and visceral atherosclerosis | CCS 114 |
| Chronic obstructive pulmonary disease and bronchiectasis | CCS 127 |
| Respiratory failure; insufficiency; arrest (adult) | CCS 131 |
| Other liver disease | CCS 151 |
| Chronic ulcer of skin | CCS 199 |
| Other injury or condition due to external causes | CCS 244 |
| Nausea and vomiting | CCS 250 |
| Delirium, dementia and amnestic and other cognitive disorders | CCS 653 |
| Developmental disorders | CCS 654 |
| Alcohol-related disorder | CCS 660 |
| Screening and history of mental health and substance abuse | CCS 663 |

**eTable 2. Diagnoses for Epic End of Life Care Index equity evaluation, with corresponding International Classification of Diseases, 10^th^ Revision (ICD-10) codes**

| **Diagnosis** | **ICD-10 codes** |
| --- | --- |
| Advanced solid malignancy | C01, C020-024, C028-031, C039-041, C048-052, C058-062, C0680, C0689, C069, C07, C080, C081, C089-091, C098, C099, C101-103, C108-113, C118, C119, C12, C130-132, C138-140, C153-155, C158-166, C168-172, C178-189, C20, C218, C220-229, C23, C240, C241, C248-254, C257-261, C269, C300, C310-313, C318-323, C328, C329, C33, C3400-3402, C3410-3412, C342, C3430-3432, C3480-3482, C3490-3492, C384, C390, C399, C450-452, C457, C459, C50011, C50012, C50019, C50021, C50022, C50029, C50111, C50112, C50119, C50121, C50122, C50129, C50211, C50212, C50219, C50221, C50222, C50229, C50311, C50312, C50319, C50321, C50322, C50329, C50411, C50412, C50419, C50421, C50422, C50429, C50511, C50512, C50519, C50521, C50522, C50529, C50611, C50612, C50619, C50621, C50622, C50629, C50811, C50812, C50819, C50821, C50822, C50829, C50911, C50912, C50919, C50921, C50922, C50929, C530, C531, C538-543, C549, C55, C561,-563, C569, C61, C641, C642, C649, C651, C652, C659, C661, C662, C669, C670-680, C688, C689, C7400-7402, C7410-7412, C7490-7492, C7800-7802, C781, C782, C7830, C7839, C784-787, C7880, C7889, C7900-7902, C7931, C7932, C7940, C7951, C7952, C7970-7972, C7989, C799, C7A00, C7A010-7A012, C7A019-7A026, C7A029, C7A090-7A096, C7A098, C7A1, C7A8, J910 |
| Hematologic malignancy | C9010, C9012, C9100-9102, C9200, C9202, C9230, C9232, C9240, C9242, C9260, C9262, C92A0, C92A2, D613, D619 |
| Chronic obstructive pulmonary disease | J410, J411, J418, J42, J430-432, J438-441, J4481, J4489, J449 |
| Fibrotic (restrictive) lung disease | D860, D862, J632, J701, J703, J704, J8410, J84112, J84113, J8417, J84170, J84178, M0510, M0519, M3213, M3321, M3391, M3481, M3502 |
| Liver disease | I8500, I8501, I8510, I8511, I864, K702, K7030, K7031, K7040, K7041, K7210, K7211, K7290, K7291, K740, K7402, K741-745, K7460, K7469, K766, K767, K7681 |
| Neurodegenerative disease | F01C0, F01C11, F01C18, C01C2-01C4, F02C18, F02C2-02C4, F03C0, F03C11, F03C18, F03C2-03C4, G10, G111, G1110, G1221, G1225, G20, G300, G301, G308, G309, G3101, G3109, G3183 G35, G7000, G7001 |
| Heart failure | I501, I5020-5023, I5030-5033, I5040-5043, I50810-50814, I5082, I5084, I5089, I509 |
| Chronic kidney disease | N184-186, N19 |
